# Analysis of 200,000 exome-sequenced UK Biobank subjects illustrates the contribution of rare genetic variants to hyperlipidaemia

**DOI:** 10.1101/2021.01.05.20249090

**Authors:** David Curtis

## Abstract

A few genes have previously been identified in which very rare variants can have major effects on lipid levels. Weighted burden analysis of rare variants was applied to exome sequenced UK Biobank subjects with hyperlipidaemia as the phenotype, of whom 44,050 were designated cases and 156,578 controls, with the strength of association characterised by the signed log 10 p value (SLP). With principal components included as covariates there was a tendency for genes on the X chromosome to produce strongly negative SLPs, and this was found to be due to the fact that rare X chromosome variants were identified less frequently in males than females. The test performed well when both principal components and sex were included as covariates and strongly implicated *LDLR* (SLP = 50.08) and *PCSK9* (SLP = -10.42) while also highlighting other genes previously found to be associated with lipid levels. Variants classified by SIFT as deleterious have on average a two-fold effect and their cumulative frequency is such that they are present in approximately 1.5% of the population. These analyses shed further light on the way that genetic variation contributes to risk of hyperlipidaemia and in particular that there are very many protein-altering variants which have on average moderate effects and whose effects can be detected when large samples of exome-sequenced subjects are available. This research has been conducted using the UK Biobank Resource.

## Introduction

We recently reported the results of analysis of 50,000 exome-sequenced UK Biobank subjects aiming to identify rare variant effects in genes influencing susceptibility to hyperlipidaemia and also briefly reviewed what was known to date about the genetic contributors to this phenotype (Curtis, 2020). The potential advantage of studying rare variants is that they have more profound, readily interpretable impacts on biology than common variants, whose effect sizes tend to be constrained by selection pressures. Rare variants with a large dominant effect in *LDLR, APOB* and *PCSK9* cause 40% of cases of familial hyperlipidaemia and there are also common variants which exert small effects on hyperlipidaemia risk (Natarajan et al., 2018; Sharifi et al., 2017; Willer et al., 2013; Wu et al., 2019). Although for most genes impaired function increases risk, the *PCSK9* variants which cause familial hyperlipidaemia produce a gain of function whereas loss of function variants cause hypobetalipoproteinemia and PCSK9 inhibitors are used as treatments to lower cholesterol levels (Dron and Hegele, 2016).

The previous analysis of 50,000 UK Biobank identified one gene, *HUWE1*, which met criteria for statistical significance after correction for multiple testing and in which there was an excess of rare and/or damaging variants in controls, suggesting that impaired functioning of this gene was protective against hyperlipidaemia. A number of other genes which were individually significant with uncorrected p<0.001, were arguably of potential interest, including *LDLR*, and, in an analysis of sets of genes, the GO gene set GENERATION OF PRECURSOR METABOLITES AND ENERGY was statistically significant. The whole UK Biobank sample consists of 500,000 subjects and a new release of data means that there is now exome sequence data available for 200,000 of them (Szustakowski et al., 2020). We report here the results of analysis of this larger dataset, which includes the original 50,000.

Large samples of exome sequenced subjects have only become available relatively recently and controversy remains about the optimal methods of analysis. Sequencing reveals very large numbers of genetic variants, many of which will have no biological effect and/or will be extremely rare, occurring in only a handful of subjects or just as singletons. The rarity of individual variants means that they need to be grouped together in a burden analysis and it is common practice to combine all variants which are predicted to completely disrupt the working of a gene, comprising: variants which introduce a stop codon; small insertions and deletions which are not a multiple of three bases and hence disrupt the amino acid code, termed frameshift variants; variants changing essential splice site sequences at intron-exon boundaries, disrupting normal splicing of exons. These three types of variant are predicted to all have a broadly similar effect no matter where they occur in the gene, consisting of a complete failure of the gene to produce normal product, and they may be referred to as loss of function (LOF) variants. It may then become possible to implicate a gene in the pathogenesis of phenotype by observing a general excess of LOF variants in that gene among cases relative to controls (Singh et al., 2016). However it is certainly the case that other kinds of variant can also cause disease. A variant which changes a codon so that it codes for a different amino acid, termed a non-synonymous variant, may alter the structure or function of the protein product in a way which dramatically affects risk but alternatively a protein altering variant may have no effect at all. The impact of a non-synonymous variant will depend crucially on the nature of the amino acid change and its position in the protein and it remains a challenging task to predict the biological effect although commonly used software such as PolyPhen and SIFT attempt this (Adzhubei et al., 2013; Kumar et al., 2009). PolyPhen designates some variants as “possibly damaging” or “probably damaging” and SIFT designates some variants as “deleterious” but different prediction programs do not always agree with each other. Nonsynonymous variants are much more frequent than LOF variants and so it would be desirable to incorporate them into burden analyses but there is a risk that doing so may simply introduce additional noise. Identifying which specific variants are most likely to have biological effects could increase power to implicate risk genes but remains a challenging task. Even variants which do not change amino acid sequence, including synonymous and intronic variants, can through various mechanisms occasionally have effects on risk and so could potentially be included.

The approach we have taken to address these issues is to carry out weighted burden analyses, in which variants judged *a priori* to be most likely to have important effects are accorded higher weights. Since selection pressures mean that common variants are unlikely to have large effects, variants are also weighted according to rarity and the detailed scheme for doing this is described in the Methods section. However a weakness of this approach to date has been that there has been little empirical evidence to inform the exact weighting scheme which would be optimal. An advantage of the large UK Biobank dataset is that it allows some exploration of the relative average effect sizes of different categories of variant and this was carried out using multivariate analyses of variant categories in addition to standard weighted burden analyses of genes and gene sets. These investigations were applied to the previously used hyperlipidaemia phenotype, defined as subjects with a diagnosis of hyperlipidaemia and/or taking cholesterol-lowering medication.

## Methods

The UK Biobank dataset was downloaded along with the variant call files for 200,632 subjects who had undergone exome-sequencing and genotyping by the UK Biobank Exome Sequencing Consortium using the GRCh38 assembly with coverage 20X at 95.6% of sites on average (Szustakowski et al., 2020). UK Biobank had obtained ethics approval from the North West Multi-centre Research Ethics Committee which covers the UK (approval number: 11/NW/0382) and had obtained informed consent from all participants. The UK Biobank approved an application for use of the data (ID 51119) and ethics approval for the analyses was obtained from the UCL Research Ethics Committee (11527/001). All variants were annotated using the standard software packages VEP, PolyPhen and SIFT (Adzhubei et al., 2013; Kumar et al., 2009; McLaren et al., 2016). To obtain population principal components reflecting ancestry, version 2.0 of *plink* (https://www.cog-genomics.org/plink/2.0/) was run with the options *--maf 0*.*1 --pca 20 approx* (Chang et al., 2015; Galinsky et al., 2016).

The hyperlipidaemia phenotype was determined in the same way as previously from four sources in the dataset: self-reported high cholesterol; reporting taking cholesterol lowering medication; reporting taking a named statin; having an ICD10 diagnosis for hyperlipidaemia in hospital records or as a cause of death (Curtis, 2020). Subjects in any of these categories were deemed to be cases with hyperlipidaemia while all other subjects were taken to be controls.

The method of analysis was the same as used previously on the smaller sample. The SCOREASSOC program was used to carry out a weighted burden analysis to test whether, in each gene, sequence variants which were rarer and/or predicted to have more severe functional effects occurred more commonly in cases than controls. Attention was restricted to rare variants with minor allele frequency (MAF) <= 0.01 in both cases and controls. As previously described, variants were weighted by overall MAF so that variants with MAF=0.01 were given a weight of 1 while very rare variants with MAF close to zero were given a weight of 10 (Curtis, 2021). Variants were also weighted according to their functional annotation using the GENEVARASSOC program, which was used to generate input files for weighted burden analysis by SCOREASSOC (Curtis, 2016, 2012). A maximum weight of 40 was assigned to variants predicted to cause complete LOF of the gene, namely stop-gained, frameshift and essential splice site variants. Other types of variant were assigned intermediate weights, for example, a weight of 5 was assigned for a synonymous variant, 10 for a non-synonymous variant and 15 for inframe insertions and deletions. Additionally, 10 was added to the weight if the PolyPhen annotation was possibly or probably damaging and also if the SIFT annotation was deleterious, meaning that a non-synonymous variant annotated as both damaging and deleterious would be assigned an overall weight of 30. In order to allow exploration of the effects of different types of variant on disease risk the variants were also grouped into broader categories to be used in multivariate analyses as described below. The full set of weights and categories is displayed in Table 1.

**Table 1.** The table shows the weight which was assigned to each type of variant as annotated by VEP, Polyphen and SIFT as well as the broad categories which were used for multivariate analyses of variant effects (Adzhubei et al., 2013; Kumar et al., 2009; McLaren et al., 2016).

| VEP / SIFT / Polyphen annotation | Weight | Category |
| --- | --- | --- |
| intergenic_variant | 1 | Unused |
| feature_truncation | 3 | Intronic, etc. |
| regulatory_region_variant | 3 | Intronic, etc. |
| feature_elongation | 3 | Intronic, etc. |
| regulatory_region_amplification | 3 | Intronic, etc. |
| regulatory_region_ablation | 3 | Intronic, etc. |
| TF_binding_site_variant | 3 | Intronic, etc. |
| TFBS_amplification | 3 | Intronic, etc. |
| TFBS_ablation | 3 | Intronic, etc. |
| downstream_gene_variant | 3 | Intronic, etc. |
| upstream_gene_variant | 3 | Intronic, etc. |
| non_coding_transcript_variant | 3 | Intronic, etc. |
| NMD_transcript_variant | 3 | Intronic, etc. |
| intron_variant | 3 | Intronic, etc. |
| non_coding_transcript_exon_variant | 3 | Intronic, etc. |
| 3_prime_UTR_variant | 10 | 3 prime UTR |
| 5_prime_UTR_variant | 5 | 5 prime UTR |
| mature_miRNA_variant | 5 | Unused |
| coding_sequence_variant | 5 | Unused |
| synonymous_variant | 5 | Synonymous |
| stop_retained_variant | 5 | Unused |
| incomplete_terminal_codon_variant | 5 | Unused |
| splice_region_variant | 5 | Splice region |
| protein_altering_variant | 10 | Protein altering |
| missense_variant | 10 | Protein altering |
| inframe_deletion | 15 | InDel, etc |
| inframe_insertion | 15 | InDel, etc |
| transcript_amplification | 15 | InDel, etc |
| start_lost | 30 | Unused |
| stop_lost | 30 | Unused |
| frameshift_variant | 40 | Disruptive |
| stop_gained | 40 | Disruptive |
| splice_donor_variant | 40 | Splice site variant |
| splice_acceptor_variant | 40 | Splice site variant |
| transcript_ablation | 20 | Disruptive |
| SIFT deleterious | 10 | Deleterious |
| PolyPhen possibly damaging | 10 | Possibly damaging |
| PolyPhen probably damaging | 10 | Probably damaging |

As described previously, the weight due to MAF and the weight due to functional annotation were multiplied together to provide an overall weight for each variant. Variants were excluded if there were more than 10% of genotypes missing in the controls or if the heterozygote count was smaller than both homozygote counts in the controls. If a subject was not genotyped for a variant then they were assigned the subject-wise average score for that variant. For each subject a gene-wise weighted burden score was derived as the sum of the variant-wise weights, each multiplied by the number of alleles of the variant which the given subject possessed. For variants on the X chromosome, hemizygous males were treated as homozygotes.

For each gene, a ridge regression analysis was carried out with lamda=1 to test whether the gene-wise variant burden score was associated with the hyperlipidaemia phenotype. To do this, SCOREASSOC first calculates the likelihood for the phenotypes as predicted by the first 20 population principal components and then calculates the likelihood using a model which additionally incorporates the gene-wise burden scores. It then carries out a likelihood ratio test assuming that twice the natural log of the likelihood ratio follows a chi-squared distribution with one degree of freedom to produce a p value. The statistical significance is summarised as a signed log p value (SLP) which is the log base 10 of the p value given a positive sign if the score is higher in cases and negative if it is higher in controls. In previous analyses it appeared that incorporating population principal components in this way satisfactorily controlled for test statistic inflation when applied to the ancestrally heterogeneous UK Biobank dataset (Curtis, 2021). However preliminary analyses of this new, larger dataset revealed that there was a slight tendency for more rare variants in X chromosome genes to be identified in females rather than males. Hence, sex was also included as a covariate along with the principal components and this produced a well-behaved test statistic, as detailed in the Results section.

Gene set analyses were carried out as before using the 1454 “all GO gene sets, gene symbols” pathways as listed in the file *c5*.*all*.*v5*.*0*.*symbols*.*gmt* downloaded from the Molecular Signatures Database at http://www.broadinstitute.org/gsea/msigdb/collections.jsp (Subramanian et al., 2005). For each set of genes, the natural logs of the gene-wise p values were summed according to Fisher’s method to produce a chi-squared statistic with degrees of freedom equal to twice the number of genes in the set. The p value associated with this chi-squared statistic was expressed as a minus log10 p (MLP) as a test of association of the set with the hyperlipidaemia phenotype.

For selected genes, additional analyses were carried out to clarify the contribution of different categories of variant. To do this, each category as listed in Table 1 was assigned a weight consisting of a different power of 10 and then GENEVARASSOC and SCOREASSOC were used to obtain scores for each subject as the sum of these weights. This allowed the overall number of variants of each category possessed by a subject to be coded as a decimal number so that, for example, a score of 1000302 would indicate that the subject possessed one of one category of variant, three of another category and two of a third category. Code was written in R to read in these scores and parse them to obtain the subject-wise counts for each category of variant (R Core Team, 2014). These were then entered into a logistic regression analysis of case-control status along with principal components and sex in order to estimate the relative contributions of different variant categories to the phenotype. The odds ratios associated with the category were estimated along with their standard errors and the Wald statistic was used to obtain a p value, except for categories in which variants occurred fewer than 50 times in which case Fisher’s exact test was applied to the raw variant counts without including covariates. The associated p value was converted to an SLP, again with the sign being positive if the mean count was higher in cases than controls.

## Results

### Results of gene-wise weighted burden tests

There were 44,054 cases with a diagnosis of hyperlipidaemia and/or taking cholesterol-lowering medication and 156,578 controls. There were 22,642 genes for which there were qualifying variants and preliminary analyses showed that there was a bias towards producing strongly negative SLPs, which was confined to genes on the X chromosome. The analyses were repeated using sex as a phenotype and this confirmed that the frequency of rare, damaging variants was higher in females for genes on the X chromosome. This would occur if the genotype calling algorithm were slightly more likely to call a female as heterozygous than a male as hemizygous. Since the frequency of cases is lower in females, the overall effect is to observe an excess of rare, damaging variants in controls rather than cases for genes on the X chromosome. Therefore the analyses were repeated for hyperlipidaemia using sex as a covariate as well as the principal components. When this was done only two genes produced strongly positive or negative SLPs, *LDLR* (SLP = 50.08) and *PCSK9* (SLP = - 10.42). The quantile-quantile (QQ) plot for the SLPs obtained for each of the remaining genes is shown in Figure 1. This shows that the test appears to be well-behaved and conforms fairly well with the expected distribution. Omitting the genes with the 100 highest and 100 lowest SLPs, which might be capturing a real biological effect, the gradient for positive SLPs is 1.08 with intercept at -0.019 and the gradient for negative SLPs is 1.04 with intercept at -0.013, indicating only modest inflation of the test statistic.

**Figure 1.**
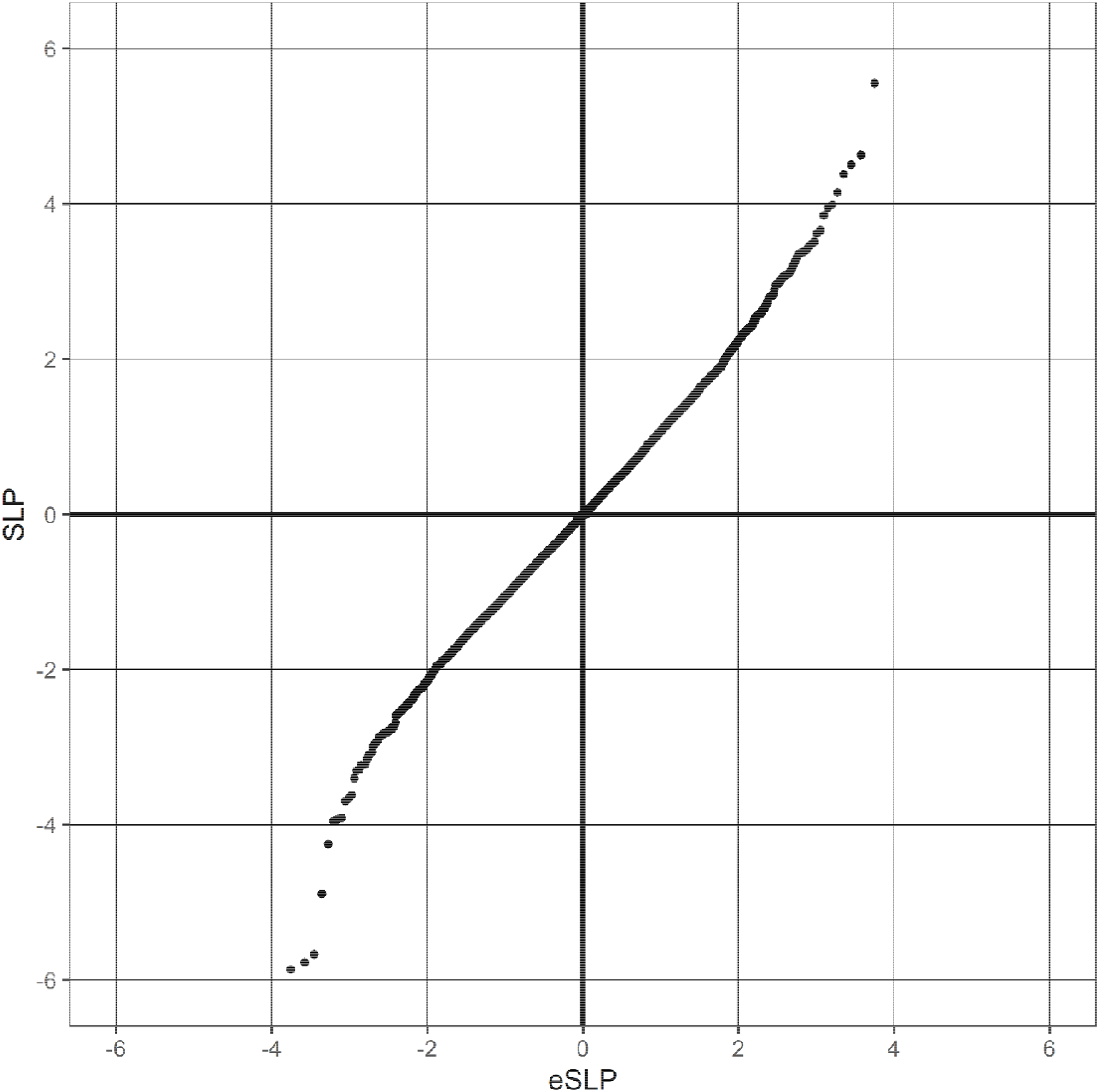
QQ plot of SLPs obtained for weighted burden analysis of association with hyperlipidaemia showing observed against expected SLP for each gene, omitting results for *LDLR* and *PCSK9*.

The role of very rare variants in both *LDLR* and *PCSK9* in the pathogenesis of familial hyperlipidaemia is already well established. However the results from the current analysis implicate a larger number of variants in these genes having a range of effects on risk of hyperlipidaemia in the population more generally. These are presented in detail below in the description of the results of the analysis of effects of different variant categories.

Given that there were 22,642 informative genes, the critical threshold for the absolute value of the SLP to declare a result as formally statistically significant is -log10(0.05/22642) = 5.66 and this was achieved by three other genes, *ANGPTL3* (SLP = -5.67), *LOC102723729* (SLP = -5.77) and *IFITM5* (SLP = -5.86). Loss of function variants in *ANGPTL3* have previously been shown to cause combined hypolipidaemia and it is the target of evinacumab, a human monoclonal antibody designed to treat hypercholesterolaemia (Doggrell, 2020; Wang and Musunuru, 2019). However *IFITM5* and *LOC102723729* do not seem to be biologically plausible candidates. *IFITM5* is involved in bone mineralisation and variants in it are a known cause of osteogenesis imperfecta (Whyte et al., 2020). *LOC102723729* is a poorly characterised lncRNA which may act as a tumour suppressor in non-small cell lung cancer (Yang et al., 2019). A total of 55 genes had SLP with absolute value of 3 or more (equivalent to uncorrected p = 0.001), whereas one would only expect around 22642/1000 = 23 by chance so a number of these may in fact be exerting some effect on risk. These are listed in Table 2 and some appear to be of particular interest and are discussed briefly as below. The results for all genes are presented in Supplementary Table 1.

**Table 2.** Genes with absolute value of SLP exceeding 3 or more (equivalent to p<0.001) for test of association of weighted burden score with hyperlipidaemia.

| Symbol | SLP | Name |
| --- | --- | --- |
| *LDLR* | 50.08 | Low Density Lipoprotein Receptor |
| *G6PC* | 5.55 | Glucose-6-Phosphatase Catalytic Subunit |
| *SULT1E1* | 4.63 | Sulfotransferase Family 1E Member 1 |
| *LOC101928415* | 4.50 | Uncharacterized LOC101928415 |
| *SLC35G1* | 4.38 | Solute Carrier Family 35 Member G1 |
| *PLA2G5* | 4.15 | Phospholipase A2 Group V |
| *CMTM7* | 3.99 | CKLF Like MARVEL Transmembrane Domain Containing 7 |
| *MIR6716* | 3.95 | MicroRNA 6716 |
| *COL4A2-AS2* | 3.85 | COL4A2 Antisense 2 |
| *DEFB131A* | 3.66 | Defensin Beta 131A |
| *OTULIN* | 3.62 | OTU Deubiquitinase With Linear Linkage Specificity |
| *FAM122C* | 3.51 | Family With Sequence Similarity 122C |
| *CMIP* | 3.48 | C-Maf Inducing Protein |
| *EIF4B* | 3.45 | Eukaryotic Translation Initiation Factor 4B |
| *PPP2R3B* | 3.41 | Protein Phosphatase 2 Regulatory Subunit B''Beta |
| *HNRNPAB* | 3.38 | Heterogeneous Nuclear Ribonucleoprotein A/B |
| *PREB* | 3.37 | Prolactin Regulatory Element Binding |
| *PEX12* | 3.36 | Peroxisomal Biogenesis Factor 12 |
| *FAM167A* | 3.35 | Family With Sequence Similarity 167 Member A |
| *ABCG5* | 3.31 | ATP Binding Cassette Subfamily G Member 5 |
| *ABCD1* | 3.26 | ATP Binding Cassette Subfamily D Member 1 |
| *PRAF2* | 3.21 | PRA1 Domain Family Member 2 |
| *CTHRC1* | 3.16 | Collagen Triple Helix Repeat Containing 1 |
| *SLC25A37* | 3.14 | Solute Carrier Family 25 Member 37 |
| *CT62* | 3.11 | Cancer/Testis Associated 62 |
| *L1TD1* | 3.09 | LINE1 Type Transposase Domain Containing 1 |
| *PIK3R6* | 3.09 | Phosphoinositide-3-Kinase Regulatory Subunit 6 |
| *FOXO3B* | 3.08 | Forkhead Box O3B |
| *FAM47A* | 3.06 | Family With Sequence Similarity 47 Member A |
| *MIR6806* | 3.06 | MicroRNA 6806 |
| *GCK* | 3.04 | Glucokinase |
| *MAPKAPK2* | 3.02 | MAPK Activated Protein Kinase 2 |
| *HLA-A* | 3.01 | Major Histocompatibility Complex, Class I, A |
| *CRYZL1* | -3.06 | Crystallin Zeta Like 1 |
| *APPBP2* | -3.08 | Amyloid Beta Precursor Protein Binding Protein 2 |
| *GFPT1* | -3.10 | Glutamine--Fructose-6-Phosphate Transaminase 1 |
| *TTR* | -3.16 | Transthyretin |
| *TXNL4A* | -3.22 | Thioredoxin Like 4A |
| *ITM2B* | -3.23 | Integral Membrane Protein 2B |
| *UBR4* | -3.23 | Ubiquitin Protein Ligase E3 Component N-Recognin 4 |
| *SV2B* | -3.29 | Synaptic Vesicle Glycoprotein 2B |
| *LOC101929609* | -3.30 | Uncharacterized LOC101929609 |
| *LOC105377994* | -3.40 | Uncharacterized LOC105377994 |
| *SNX17* | -3.62 | Sorting Nexin 17 |
| *ANGPTL4* | -3.66 | Angiopoietin Like 4 |
| *NPC1L1* | -3.70 | NPC1 Like Intracellular Cholesterol Transporter 1 |
| *CTXN2* | -3.91 | Cortexin 2 |
| *TBC1D8* | -3.93 | TBC1 Domain Family Member 8 |
| *LOC107985474* | -3.95 | Uncharacterized LOC107985474 |
| *PPP1R3G* | -4.25 | Protein Phosphatase 1 Regulatory Subunit 3G |
| *APOC3* | -4.89 | Apolipoprotein C3 |
| *ANGPTL3* | -5.67 | Angiopoietin Like 3 |
| *LOC102723729* | -5.77 | Uncharacterized LOC102723729 |
| *IFITM5* | -5.86 | Interferon Induced Transmembrane Protein 5 |
| *PCSK9* | -10.42 | Proprotein Convertase Subtilisin/Kexin Type 9 |

*G6PC* (SLP = 5.55) is of interest because mutations acting recessively cause glycogen storage disease type I (GSD1, von Gierke disease, incidence ∼1/100,000), which includes hyperlipidaemia as part of the phenotype (Kishnani et al., 2014). Rare homozygous variants in *ABCG5* (SLP = 3.31) can produce sitosterolaemia and are a known cause of homozygous familial hypercholesterolaemia (Cuchel et al., 2014). Mutations in *ABCD1* (SLP = 3.26) cause X-linked adrenoleukodystrophy, which results in elevated levels of very long chain fatty acids in plasma and tissues (Engelen et al., 2012). Variants in *GCK* (SLP = 3.04) are known to cause maturity-onset diabetes of the young (MODY) with mild hyperglycaemia and lower triglyceride levels than other forms of type 2 diabetes (Ma et al., 2019). Like *ANGPTL3, ANGPTL4* (SLP = -3.66) modulates the activity of lipoprotein lipase (LPL) and inactivating variants in it have previously been shown to be associated with hypolipidaemia (Dron and Hegele, 2016). The product of *NPC1L1* (SLP = -3.70) is essential for intestinal sterol absorption and is the molecular target of ezetimibe, a potent cholesterol absorption inhibitor that lowers blood cholesterol (Betters and Yu, 2010). Homozygous knockout of *PPP1R3G* (SLP = -4.25) mitigates high-fat diet induced obesity in mice (Zhang et al., 2017). Variants in *APOC3* (SLP = -4.89) have previously shown to be protective against hyperlipidaemia risk (Dron and Hegele, 2016).

While two ATP binding cassette transporter genes, *ABCG5* and *ABCD1*, produced SLPs above 3, a third, *ABCA1* (SLP = -2.91), was only marginally less significant. The product of *ABCA1* is responsible for transporting cholesterol out of cells and homozygous or compound heterozygous variants in it cause Tangier disease, a familial HDL deficiency syndrome, while heterozygous variants are associated with reduced HDL levels (Maranghi et al., 2019; Puntoni et al., 2012). It has an established role in the regulation of HDL and there are reports that common variants in it are associated with plasma lipid levels (Koldamova et al., 2014; Lu et al., 2018).

### Results of gene set analyses

An initial run of the gene set analyses tended to highlight sets containing hundreds of genes which included one or more of the genes with absolute SLPs over 3 as listed in Table 2 so the analyses were repeated with these genes and *ABCA1* omitted to see if any additional genes of interest could be identified. Given that 1,454 sets were tested a critical MLP to achieve to declare results significant after correction for multiple testing would be log10(1454*20) = 4.46 and this was not achieved by any set. Inspection of the results for the highest scoring sets did not reveal any additional genes which might obviously be involved in hyperlipidaemia risk. The results for all sets are provided in Supplementary Table 2.

### Results of variant category analyses

For the two genes showing the most definite evidence of association, *LDLR* and *PCSK9*, a logistic regression analysis of different categories of variant was carried out to elucidate their relative contributions. The results for *LDLR* are shown in Table 3A. It can be seen that disruptive variants, comprising stop variants and frameshift variants, are significantly associated with caseness (SLP = 16.95) with a large effect on risk (OR = 40.02 (11.83 - 135.33)). There were 34 of these variants, of which only 3 were seen in controls. Essential splice site variants also exerted a large effect on risk (OR = 10.4 (1.9 - 56.7)), SLP = 5.55, with 11 out of 13 being seen in cases. Stop variants, frameshift variants and essential splice variants are expected to cause LOF but the results show that other variants which do not severely disrupt the gene but which produce changes in amino acid sequence also have moderate effects on risk. There were 6,747 nonsynonymous variants and this category was associated with OR = 1.15 (1.05 - 1.25). However of these 1,175 were annotated by SIFT as “deleterious” and this category has OR = 1.74 (1.41 - 2.14) while the risk associated with an annotation by PolyPhen of “probably damaging” was smaller, 1.30 (1.03 - 1.65), and there was no significant risk associated with an annotation of “possibly damaging”. Inframe insertion/deletion variants were observed on 10 occasions and detailed inspection of the results revealed that these consisted of deletions at 4 different positions, one of which occurred in 7 different subjects. All 10 of the subjects with one of these deletions was a case (SLP=6.58). By contrast, genetic variants which did not affect protein sequence in general did not have significant effects on risk. The exception was that the “Splice Region” category seemed to exert a protective effect, with OR = 0.86 (0.81 - 0.92), SLP = -5.21. This was driven by rs72658867, which had frequency 0.012 in controls and 0.0097 in cases and which has been previously reported to be associated with lower cholesterol and lower risk of coronary artery disease (Gretarsdottir et al., 2015). When the analysis was repeated with this variant removed, there was no general tendency for splice region variants to be associated with risk (OR = 1.13 (0.99 - 1.29), SLP = 1.20).

**Table 3.**
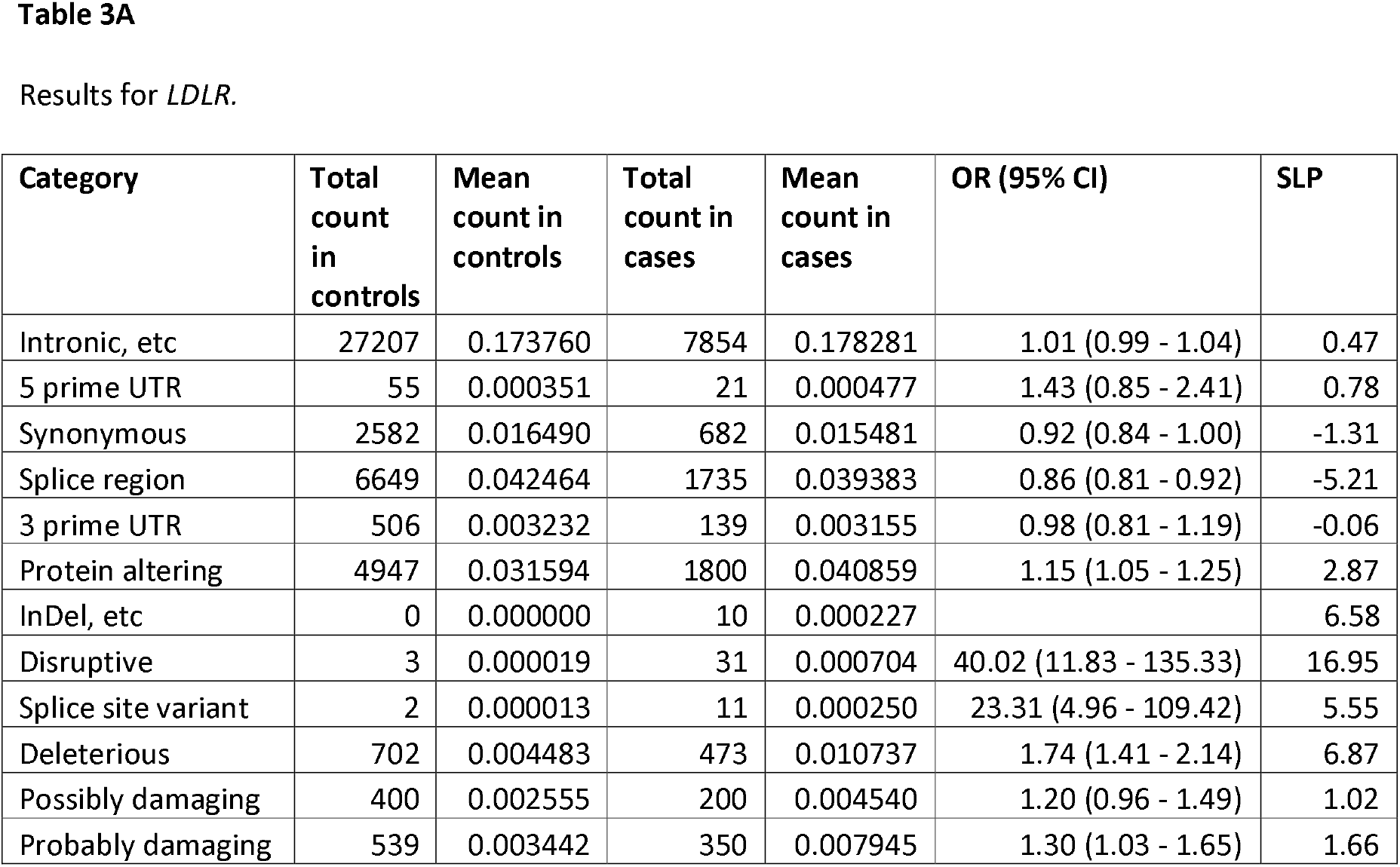
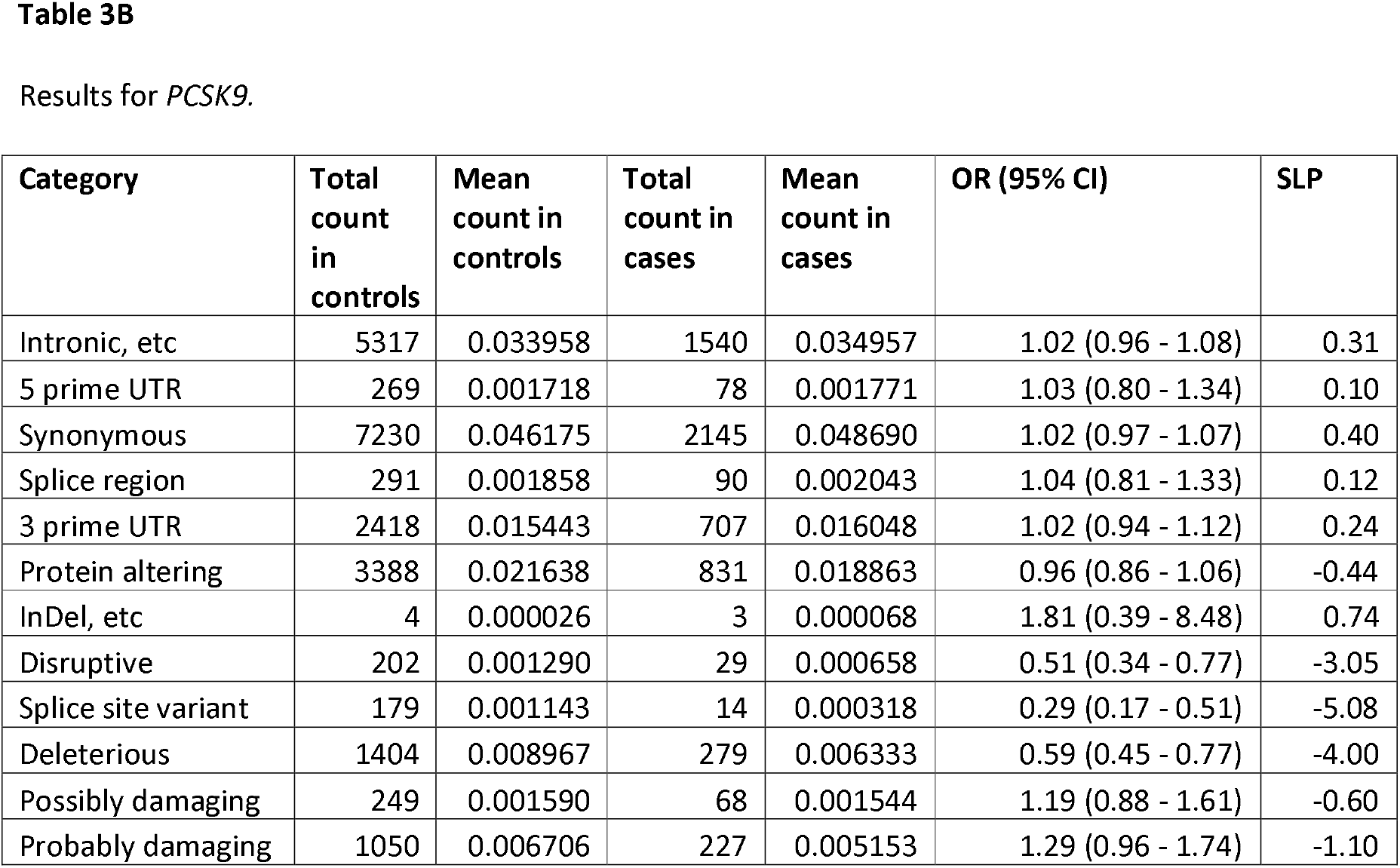
Results from logistic regression analysis showing the contribution different categories of variant within a gene make to risk of hyperlipidaemia. Odds ratios for each category are estimated including principal components and sex as covariates.

| Category | Total count in controls | Mean count in controls | Total count in cases | Mean count in cases | OR (95% CI) | SLP |
| --- | --- | --- | --- | --- | --- | --- |
| Intronic, etc | 27207 | 0.173760 | 7854 | 0.178281 | 1.01 (0.99 - 1.04) | 0.47 |
| 5 prime UTR | 55 | 0.000351 | 21 | 0.000477 | 1.43 (0.85 - 2.41) | 0.78 |
| Synonymous | 2582 | 0.016490 | 682 | 0.015481 | 0.92 (0.84 - 1.00) | -1.31 |
| Splice region | 6649 | 0.042464 | 1735 | 0.039383 | 0.86 (0.81 - 0.92) | -5.21 |
| 3 prime UTR | 506 | 0.003232 | 139 | 0.003155 | 0.98 (0.81 - 1.19) | -0.06 |
| Protein altering | 4947 | 0.031594 | 1800 | 0.040859 | 1.15 (1.05 - 1.25) | 2.87 |
| InDel, etc | 0 | 0.000000 | 10 | 0.000227 |  | 6.58 |
| Disruptive | 3 | 0.000019 | 31 | 0.000704 | 40.02 (11.83 - 135.33) | 16.95 |
| Splice site variant | 2 | 0.000013 | 11 | 0.000250 | 23.31 (4.96 - 109.42) | 5.55 |
| Deleterious | 702 | 0.004483 | 473 | 0.010737 | 1.74 (1.41 - 2.14) | 6.87 |
| Possibly damaging | 400 | 0.002555 | 200 | 0.004540 | 1.20 (0.96 - 1.49) | 1.02 |
| Probably damaging | 539 | 0.003442 | 350 | 0.007945 | 1.30 (1.03 - 1.65) | 1.66 |
| Intronic, etc | 5317 | 0.033958 | 1540 | 0.034957 | 1.02 (0.96 - 1.08) | 0.31 |
| 5 prime UTR | 269 | 0.001718 | 78 | 0.001771 | 1.03 (0.80 - 1.34) | 0.10 |
| Synonymous | 7230 | 0.046175 | 2145 | 0.048690 | 1.02 (0.97 - 1.07) | 0.40 |
| Splice region | 291 | 0.001858 | 90 | 0.002043 | 1.04 (0.81 - 1.33) | 0.12 |
| 3 prime UTR | 2418 | 0.015443 | 707 | 0.016048 | 1.02 (0.94 - 1.12) | 0.24 |
| Protein altering | 3388 | 0.021638 | 831 | 0.018863 | 0.96 (0.86 - 1.06) | -0.44 |
| InDel, etc | 4 | 0.000026 | 3 | 0.000068 | 1.81 (0.39 - 8.48) | 0.74 |
| Disruptive | 202 | 0.001290 | 29 | 0.000658 | 0.51 (0.34 - 0.77) | -3.05 |
| Splice site variant | 179 | 0.001143 | 14 | 0.000318 | 0.29 (0.17 - 0.51) | -5.08 |
| Deleterious | 1404 | 0.008967 | 279 | 0.006333 | 0.59 (0.45 - 0.77) | -4.00 |
| Possibly damaging | 249 | 0.001590 | 68 | 0.001544 | 1.19 (0.88 - 1.61) | -0.60 |
| Probably damaging | 1050 | 0.006706 | 227 | 0.005153 | 1.29 (0.96 - 1.74) | -1.10 |

Table 3B shows the results of the variant category analysis for *PCSK9*. It can be seen that these are broadly similar to those obtained for *LDLR*, albeit in the opposite direction because impaired function of *PCSK9* reduces risk of hyperlipidaemia. Disruptive, essential splice site and missense variants annotated as “deleterious” by SIFT are all significantly more common in controls and have an overall OR of around 0.5. It is interesting to note that these categories of variant occur more frequently in *PCSK9* than in *LDLR*. In *LDLR* there are only 34 disruptive variants whereas in *PCSK9* there are 291 and in *LDLR* there are only 13 essential splice site variants while in *PCSK9* there are 193.

These results allow us to gain some insight into the overall impact of variants in these genes on the risk of hyperlipidaemia in the general population. For variants in *LDLR* which are nonsynonymous and annotated as “deleterious” by SIFT, the overall estimated OR is 1.15*1.74 = 2. It should be emphasised that this estimate is for the average effect of such variants and that there is likely to be considerable variation, with some of these variants exerting marked effects on risk while others may have trivial effects or may even be protective. There are 889 of these variants and, since they are rare, few people have more than one of them so that we can say that around 850 out of the 200,000 subjects, or slightly less than 0.5%, have a deleterious variant in *LDLR* which, on average, about doubles the odds of hyperlipidaemia. This compares with the 47 LOF variants which confer high risk but which occur in only 0.02% of subjects. Deleterious variants in *PCSK9* on average have OR of about 0.5 and occur in 0.8% of subjects while LOF variants have a similar OR and occur in 0.2% of subjects. Broadly speaking, it seems that about 1.5% of people will have a rare coding variant in one of these two genes which either doubles or halves the odds of developing hyperlipidaemia.

### Results for selected genes

It is relevant also to report certain genes which produced negative results. With the exception of *LDLR*, none of the genes highlighted by the previous analysis of 50,000 UK Biobank exomes showed any evidence for association in this enlarged sample once sex was included as a covariate. These genes consist of *HUWE1, CXorf56, RBP2, STAT5B, NPFFR1, ACOT9, GK, ADIPOQ, SURF1, ADRB3, GYG2, PHKA1* and *PHKA2* (Curtis, 2020). *HUWE1* and a number of others are located on the X chromosome and with hindsight it appears that they may have produced strongly negative SLPs as a consequence of the reduced frequency of variants called on the X chromosome in males, while other results may have simply been due to chance. Other genes for which notably negative results are obtained are *APOB* (SLP = 0.00) a known cause of familial hypercholesterolaemia, and *HMGCR* (SLP = -0.07), which codes for the rate-limiting enzyme in cholesterol synthesis which is the target of statins (LaRosa et al., 1999). Also negative was *STAP1* (SLP = -1.02), for which there were initial claims of an association with familial hypercholesterolaemia although more recent work has thrown doubt on this (Kanuri et al., 2020). These three genes were also subjected to the variant category analysis and no category of variant within them showed significant association with hyperlipidaemia.

## Discussion

These analyses provide a broad overview of contributions of rare coding genetic variants to the risk of hyperlipidaemia. There are a number of issues worthy of further comment.

The observation that in this dataset rare X chromosome variants are called more frequently in females than in males is important to recognise. Unless this effect is allowed for, for example by incorporating sex as a covariate, artefactual results may be produced for any phenotype whose prevalence varies with sex. With hindsight, this occurred in the earlier analysis of the 50,000 exomes and led to the identification as some genes on the X chromosome as being potentially relevant. Going forward, researchers need to be aware of this phenomenon and deal with it appropriately.

Working with biobank datasets can pose particular challenges compared to traditional case-control studies. In a case-control study one can control recruitment and assess subjects against pre-specified criteria. With the UK Biobank one has a self-selected sample of volunteers along with information about a broad range of phenotypes but some measures are only available for a subset of the sample.

The phenotype studied here is intended to broadly capture clinically significant hyperlipidaemia, using as it does a combination of the diagnosis and the most commonly used treatments. However this phenotype clearly differs from what one might use in a more systematically assessed sample. No attempt was made to incorporate actual measures of blood lipids, in part because these might be distorted by treatment effects. Some subjects will have been prescribed statins purely on the basis of raised lipids found during routine clinical assessment whereas other subjects with somewhat lower levels might be receiving them because they had cardiovascular disease. Likewise, some subjects classified as controls might in fact have hyperlipidaemia which has not been diagnosed. Thus, the phenotype is understood to be a quite noisy and a distant consequence of the immediate biological effects of any functional genetic variants. Another issue is that the participants represent a relatively healthy group of subjects. People with severe hyperlipidaemia which had resulted in early death would not be included, meaning that the effect sizes observed in this sample may tend to be underestimates.

The current analysis highlights a number of genes for which very rare variants with large effect size have previously been shown to impact lipid levels and demonstrates that large numbers of additional variants with more moderate effect also make a broader contribution to risk in the general population. This is most clearly the case for *LDLR* and *PCSK9* but there are a few of other genes which probably also show this effect, especially *ANGPTL3* and *ANGPTL4*. Conversely, other genes which are implicated as monogenic causes of severe familial hyperlipidaemias, such as *APOB* and *STAP1*, are not identified by this approach as making broader contributions to hyperlipidaemia risk. The analyses highlight three genes which are already the targets of lipid-lowering therapies, *PCSK9, ANGPTL3* and *NPC1L1*, but completely failed to detect an effect for *HMGCR*, which encodes the target of statins. The approach used is intended to detect the additive effects of variants which are individually very rare but which cumulatively have an effect on the function of a gene. Hence it is not expected to be successful if the effect of some variants impairing gene function may be counterbalanced by others which produce a gain of function. It is necessary to group variants because when a variant is only observed in a handful of subjects it is not possible to draw firm conclusions about its effect. There might be scope to gain power by devising more sophisticated approaches to variant classification, for example related to the more specific predictions about effect on the protein product.

Estimating the effect on risk of different categories of variant within *LDLR* and *PCSK9* broadly confirms what we might have expected. LOF variants, comprising stop, frameshift and splice site variants, have large effects. Variants categorised as “deleterious” by SIFT have moderate effects on risk, whereas the categorisation as “probably damaging” by PolyPhen is associated with a somewhat smaller effect. The annotation of “possibly damaging” does not seem to have much utility in this context. The analyses show that even nonsynonymous variants in *LDLR* which do not have any of these annotations are still, on average, associated with a slightly increased risk, with OR = 1.15. Variants which do not cause changes in amino acid sequence do not in general seem to exert an appreciable effect on risk. These findings will be of use in constructing weighting schemes for future analyses of this nature. The variants are mostly all individually extremely rare but in total there are far more nonsynonymous variants than LOF variants.

The general picture which emerges is that there is a relatively small number of genes in which variants which are individually extremely rare make an appreciable contribution to the overall risk of developing hyperlipidaemia. Few variants cause LOF but those which do have a large effect, whereas far larger numbers of nonsynonymous variants tend to exert more moderate effects. Nevertheless, the cumulative frequency of these variants remains low. If we confine attention to the results about which we can feel most confident, it seems that fewer than 2% of people carry a variant which might halve or double risk. It will be possible to refine estimates such as this as more data becomes available, for example from the remaining 300,000 UK Biobank subjects for whom exome sequence data is yet to be provided. With a larger dataset it will become possible to draw more definitive conclusions about individual genes and to make more accurate estimates of effect sizes.

The availability of sequence data from a large number of subjects has allowed insights into the contribution which rare coding genetic variants can make to hyperlipidaemia, an important phenotype which is also associated with a variety of socioeconomic and environmental risk factors. More detailed analyses may focus on specific genes and/or variants, may investigate signals of selection pressures, may look at interactions between different genetic and environmental variables and may explore the development of individualised risk assessments. Hyperlipidaemia provides a useful paradigm of a common complex trait and similar approaches can be applied to other phenotypes.

## Data Availability

The raw data is available on application to UK Biobank. Detailed results with variant counts cannot be made available because they might be used for subject identification. Scripts and relevant derived variables including principle components and variant annotations will be deposited in UK Biobank.

## Conflicts of interest

The author declares he has no conflict of interest.

## Data availability

The raw data is available on application to UK Biobank. Detailed results with variant counts cannot be made available because they might be used for subject identification. Relevant derived variables including principal components and variant annotations will be deposited in UK Biobank. Scripts and software used to carry out the analyses are available at https://github.com/davenomiddlenamecurtis.

## Acknowledgments

This research has been conducted using the UK Biobank Resource. The author wishes to acknowledge the staff supporting the High Performance Computing Cluster, Computer Science Department, University College London. This work was carried out in part using resources provided by BBSRC equipment grant BB/R01356X/1.

## Notes

### Competing Interest Statement

The authors have declared no competing interest.

### Funding Statement

No external funding

### Author Declarations

UK Biobank had obtained ethics approval from the North West Multi-centre Research Ethics Committee which covers the UK (approval number: 11/NW/0382) and had obtained informed consent from all participants. The UK Biobank approved an application for use of the data (ID 51119) and ethics approval for the analyses was obtained from the UCL Research Ethics Committee (11527/001).

### Summary of Updates

Additional references and additional discussion.

